# Long-term antibody response to SARS-CoV-2 in children

**DOI:** 10.1101/2022.02.11.22270611

**Authors:** Gabor A. Dunay, Madalena Barroso, Mathias Woidy, Marta K. Danecka, Geraldine Engels, Katharina Hermann, Friederike S. Neumann, Kevin Paul, Jan Beime, Gabriele Escherich, Kristin Fehse, Lev Grinstein, Franziska Haniel, Luka J. Haupt, Laura Hecher, Torben Kehl, Christoph Kemen, Markus J. Kemper, Robin Kobbe, Aloisa Kohl, Thomas Klokow, Dominik Nörz, Jakob Olfe, Friderike Schlenker, Jessica Schmiesing, Johanna Schrum, Freya Sibbertsen, Philippe Stock, Stephan Tiede, Eik Vettorazzi, Dimitra E. Zazara, Antonia Zapf, Marc Lütgehetmann, Jun Oh, Thomas S. Mir, Ania C. Muntau, the C19.CHILD Study Group, Søren W. Gersting

## Abstract

Almost two years into the pandemic and with vaccination of children significantly lagging behind adults, long-term pediatric humoral immune responses to SARS-CoV-2 are understudied. The C19.CHILD Hamburg (COVID-19 Child Health Investigation of Latent Disease) Study is a prospective cohort study designed to identify and follow up children and their household contacts infected in the early 2020 first wave of SARS-CoV-2. We screened 6113 children <18 years by nasopharyngeal swab-PCR in a low-incidence setting after general lockdown, from May 11 to June 30, 2020. 4657 participants underwent antibody testing. Positive tests were followed up by repeated PCR and serological testing of all household contacts over 6 months. In total, the study identified 67 seropositive children (1.44 %), the median time after infection at first presentation was 83 days post-symptom onset (PSO). Follow up of household contacts showed incomplete seroconversion in most families, with higher seroconversion rates in families with adult index cases compared to pediatric index cases (OR: 1.79, P=0.047). Most importantly, children showed sustained seroconversion up to nine months PSO, and serum antibody concentrations persistently surpassed adult levels (ratio serum IgG Spike children vs. adults 90 days PSO: 1.75, P<0.001, 180 days: 1.38, P=0.01, 270 days: 1.54, P=0.001).

In a low-incidence setting, SARS-CoV-2 infection and humoral immune response present distinct patterns in children including higher antibody levels, and lower seroconversion rates in families with pediatric index cases. Children show long-term SARS-CoV-2 antibody responses. These findings are relevant to novel variants with increased disease burden in children, as well as for the planning of age-appropriate vaccination strategies.

## Introduction

The SARS-CoV-2 pandemic has been responsible for over five million deaths [1]. Severe disease due to acute COVID-19 is rare in children and teenagers [2–4], and the pediatric inflammatory multisystem syndrome (PIMS, also termed MIS-C) attributable to SARS-CoV-2 only occurs in a small minority of cases [5,6]. Vaccination of adults and teenagers is now widespread even though vaccination rates show large regional differences [1]. Vaccination of younger children above the age of 5 years [7] has been ongoing for several months (US) to several weeks (Europe) and even longer in China at the time of this report. Emerging viral variants, especially the most recent omicron variant (B.1.1.52) appear to escape neutralizing antibodies provided by vaccination [8]. Furthermore, the pediatric population may be at increased risk, because initial reports of the omicron variant suggest an increased hospitalization rate in children compared to earlier variants [9].

Despite the pressing need to better understand long-term antibody responses in children, longitudinal studies over longer periods after natural infection are almost completely lacking in this age group. Here, we present a prospective cohort study of 67 convalescent children and their household contacts including a quantitative follow up of antibody responses in household clusters up to nine months after symptomatic infection. The children were recruited in the C19.CHILD Hamburg Study (COVID-19 Child Health Investigation of Latent Disease), in which over six thousand children under 18 years of age were screened for acute SARS-CoV-2 infection by PCR and 4657 serological tests were performed. The recruitment to the study followed a general lockdown after the initial wave of ancestral SARS-CoV-2 infection in 2020.

## Methods

### Study population and ethical approval

We conducted an observational prospective multi-center cohort study of children under eighteen years of age in all five pediatric hospitals of the Free and Hanseatic City of Hamburg, Germany; AKK Altonaer Kinderkrankenhaus, Helios Mariahilf Kinderklinik, Klinik für Kinder-und Jugendmedizin der Asklepios Klinik Nord-Heidberg, Katholisches Kinderkrankenhaus Wilhelmstift, and Werner und Michael Otto Universitätskinderklinik at the University Medical Center Hamburg-Eppendorf (Kinder-UKE) from May 11 until June 30, 2020. Patients presenting to the study hospitals were invited to participate. Healthy individuals were welcomed to enroll through the C19.CHILD Study Clinic at Kinder-UKE. Filling out of a standardized questionnaire, as well as taking a nasopharyngeal swab for the PCR detection of SARS-CoV-2 RNA were obligatory for all participants during the screening phase, whereas a venous blood draw for the detection of antibodies directed against SARS-CoV-2 was optional.

Recruitment and consent procedures were equivalent in all participating hospitals. A written informed consent was obtained for each participating subject (*Supplementary Methods*). The study was approved by the local ethical committee of Hamburg (reference number: PV7336) and published at clinicaltrials.gov (NCT04534608).

Children with a positive PCR and/or antibody test, as well as their household contacts (referred to as family members), were invited to three follow-up appointments. The follow ups were carried out at the earliest possible timepoint within days of screening, then at three- and six months after screening at the C19.CHILD Study Clinic. Here, an extended questionnaire including a detailed history of infection was completed, and nasopharyngeal swabs and venous blood samples were collected from all family members willing to participate for SARS-CoV-2 viral RNA and antibody testing.

### Sample processing and viral RNA detection

Viral RNA was extracted from nasopharyngeal swabs using a Tecan Freedom Evo liquid handler system (Tecan) and the NucleoSpin 96 Virus Core Kit (Macherey-Nagel; refer to *Supplementary Methods* for details). For the detection of the SARS-CoV-2 RNA the RealStar SARS-CoV-2 RT-PCR Kit 1.0 (Altona Diagnostics) was used according to the manufacturer’s recommendations. The dual target assay detects the viral E-gene (B-βCoV specific) and S-gene (SARS-CoV-2 specific). Samples with positivity in both gene targets were considered positive. SARS-CoV-2 RNA detection of the follow-up participants was performed using the ECDC recommended E-gene assay [10] adapted for high-throughput PCR (Cobas 6800, Roche), as described by S. Pfefferle *et al*. [11].

### Antibody measurements

SARS-CoV-2-specific antibodies (IgA/IgM/IgG) against the viral nucleocapsid protein were detected in sample serum using the Elecsys® Anti-SARS-CoV-2 Ig assay (Roche) on the cobas e411 system (Roche). Positive results were confirmed in the same serum sample with the LIAISON® SARS-CoV-2 IgG serology test (DiaSorin). This alternative assay allows for quantification of SARS-CoV-2 antibodies (IgG) against the S1 and S2 subunits of the spike protein. Assays were performed IVD conform according to the manufacturer’s recommendations including thresholds for positivity (see *Supplementary Methods*). Following the guidelines of the CDC for a low prevalence setting (<5 %) [12], an orthogonal testing algorithm was used, therefore samples were only considered seropositive if positive results were obtained with both assays.

### Statistical Analysis

Frequency of demographic and clinical features were reported for study subjects stratified according to SARS-CoV-2 test results or age groups. Age is shown as mean ± standard deviation, and categorical variables as counts and/or percentages. The primary aim was to estimate the prevalence of SARS-CoV-2 in children and the sample size was justified in this respect. Since only one child was acutely infected, this primary question cannot be answered. Therefore, all analyses are secondary and explorative. Accordingly, the P-values are not adjusted for multiplicity and are only reported as descriptive measures. Children from the same family were included in the study. This creates a cluster structure that must be adequately considered in the statistical methodology. We therefore used mixed logistic models with the family as random intercept, where appropriate. Another issue is the low number of events, which can lead to separation problems and thus instable estimates. We addressed this by using penalized models provided in the R package GLMMadaptive. For the longitudinal analysis of serum antibody concentrations, a generalized linear mixed model was used with random effects for individuals nested within families. The tweedie distribution, which is implemented in the R package glmmTMB, was chosen in this model to account for measurements below the detection limit and additionally, a zero-inflation was allowed to reflect negative test results. The results were robust with respect to the link function, which was verified in sensitivity analyses. Analyses were performed using R software (version 4.0.2) and GraphPad Prism (version 8.4).

## Results

We screened 6113 children under eighteen years of age in the Free and Hanseatic City of Hamburg, Germany (population 1,845,017 on April 30, 2020) within a defined time period (May 11 – June 30, 2020) and included 5908 study participants from 4506 families in the C19.CHILD study. Children were recruited regardless of the presence or absence of any COVID-19 symptoms. The study included 3145 (53 %) volunteers recruited through the C19.CHILD Study Clinic at Kinder-UKE and 2763 (47 %) subjects who presented at one of the Hamburg pediatric hospitals for reasons independent of this study. In case of a positive PCR and/or antibody test, children were invited for follow-up tests along with all contact persons living in the same household.

One child tested PCR positive for SARS-CoV-2. Two antibody tests were used for this study. For analysis, the Roche nucleocapsid protein total IgA/IgM/IgG test was defined as screening antibody test (available for 4657 participants, 79 % of total included) and the DiaSorin S1/S2 spike IgG was defined as confirmatory test. A total of 67 children, 1.44 % of all tested, had a positive antibody reaction in both antibody tests. Participants were recalled for a second testing shortly after positive antibody screening, typically within six days (mean 5.8). Recall testing was available for 50 children, all tests remained positive as determined by both methods (*Supplementary Table S2*).

In this cohort, the likelihood of seroconversion to SARS-CoV-2 increased with age, the mean age of seropositive children was two years higher than that of seronegative children (10.3 vs. 8.3 years, P=0.001, *Table 1*). We observed a continuous age effect, where the probability of seroconversion increased with an odds ratio of 1.11 per year of age (95 % CI: 1.03-1.21, P=0.009 *Supplementary Figure 1*) in the cohort of screened children under eighteen years. Inclusion of more than one child per family created a cluster structure, which we considered by applying mixed logistic regression models.

**Table 1:** Baseline characteristics of study population. Age is given as mean (standard deviation), all other are counts (percent) or counts/N (percent) in case of missing data. Age is compared using Student’s t-test, all binary data is compared using Fisher’s exact test.

|  | Negative in either test<br>(N=4590) | Positive Roche &<br>DiaSorin (N=67) | Total (N=4657) | p-value |
| --- | --- | --- | --- | --- |
| Age (years) | 8.3 (5.0) | 10.3 (4.4) | 8.3 (5.0) | 0.001 |
| Female gender | 2172 (47.3 %) | 33 (49.3 %) | 2205 (47.3 %) | 0.806 |
| Underlying medical conditions | 1334 (29.1 %) | 16 (23.9 %) | 1350 (29.0 %) | 0.416 |
| Age-appropriate vaccination complete | 4088/4428 (92.3 %) | 65/66 (98.5 %) | 4153/4494<br>(92.4 %) | 0.060 |
| Contact to persons with known<br>SARS-CoV-2 Infection | 92/1712 (5.4 %) | 27/33 (81.8 %) | 119/1745<br>(6.8 %) | <0.001 |

Seroconversion rates did not show any gender differences in this cohort (*Table 1*). Children and adolescents recruited in the participating hospitals often suffered from a complex chronic disease (29% indicated an underlying medical condition, detailed in *Supplementary Table S1*). One might expect a high proportion of vulnerable individuals in the C19.CHILD study population regarding infection rates or severity of disease [13]. However, children with an underlying medical condition representing almost one third of the study cohort did not show a difference in the seroconversion rate (*Table 1, Supplementary Table S1*). When comparing self-reported, age-appropriate vaccinations with seroconversion status, children with a complete vaccination status were more likely to be seropositive than children with missed vaccinations (*Table 1*, 98.5 vs. 92.3 %, P=0.060). Participants and their guardians were asked to report on any persistent symptoms associated with COVID-19 in the last 14 days before screening. Notably, the only symptom that was more common in seropositive children was loss of taste (seronegatives 15/4590, 1.3 %, seropositives 3/67, 21.4 %, p=0.001).

We further looked at the effect of self-reported exposure to persons with confirmed SARS-CoV-2 infection on seroconversion rates. Exposure was reported for 119 children from whom antibody tests were also available. Twenty-seven of these children had a confirmed positive screening antibody test (22.7 %) and 20 were available for recall testing, all of whom tested positive for both nucleocapsid IgA/IgG/IgM and spike IgG. Children aged 12 - <18 years (seroconversion 48.1 %, OR: 3.2, 95 % CI: 0.5-18.9) as well children aged 6 - <12 years (seroconversion 40.7 %, OR: 1.1, 95 % CI: 0.2-5.6) had a higher probability of seroconversion than children aged 0 - <6 years (seroconversion 11.1 %, OR: 1) after contact to SARS-CoV-2. Furthermore, seroconversion was more likely to happen if the contact had been within the family (92.6 % of seropositives after contact, OR: 20.9, 95 %, CI: 3.0-143.8), than if contacts had been outside the family (7.4 % of seropositives after contact, OR: 1, *Table 2*).

**Table 2:** Impact of age and type of contact to a person infected with SARS-CoV-2 on the risk of seroconversion. Odds ratios are estimates from a multiple mixed logistic model.

|  | Negative in either test<br>(N=92) | Positive Roche & DiaSorin<br>(N=27) | Total<br>(N=119) | Odds Ratio<br>(95 % CI) |
| --- | --- | --- | --- | --- |
| Age (grouped) |  |  |  |  |
| 0 to <6 | 21 (22.8 %) | 3 (11.1 %) | 24 (20.2 %) | 1 (reference) |
| 6 to <12 | 42 (45.7 %) | 11 (40.7 %) | 53 (44.5 %) | 1.1 (0.2, 5.6) |
| 12 to <18 | 29 (31.5 %) | 13 (48.1 %) | 42 (35.3 %) | 3.2 (0.5, 18.9) |
| Type of contact |  |  |  |  |
| outside family | 60 (65.2 %) | 2 (7.4 %) | 62 (52.1 %) | 1 (reference) |
| inside family | 32 (34.8 %) | 25 (92.6 %) | 57 (47.9 %) | 20.9 (3.0, 143.8) |

Next, seroconversion among household contacts was analyzed. We recalled 43 families for an initial follow-up visit within days after screening (median 83 days post symptom onset, range 21-121 days). Seroconversion within families was only complete in a minority of cases (*Figure 1 A, Supplementary Table S2*), where seropositivity was again defined based on a positive result for both nucleocapsid IgA/IgG/IgM and spike IgG. Thirty-eight families were able to identify the first infected person in the household (index case) based on the beginning of symptoms based on a known contact to an infected person. Notably, when comparing seroconversion rates within individual families, we found that they were lower when the index case was younger than eighteen years than for adult index cases (mean seroconversion rate 48.9 % vs. 73.8 %, *Figure 1 B*). Applying logistic regression to model the risk of seroconversion with the type of index case as predictor confirmed these findings (OR: 1.79, 95 % CI: 1.01-3.19, P=0.047).

**Figure 1:**
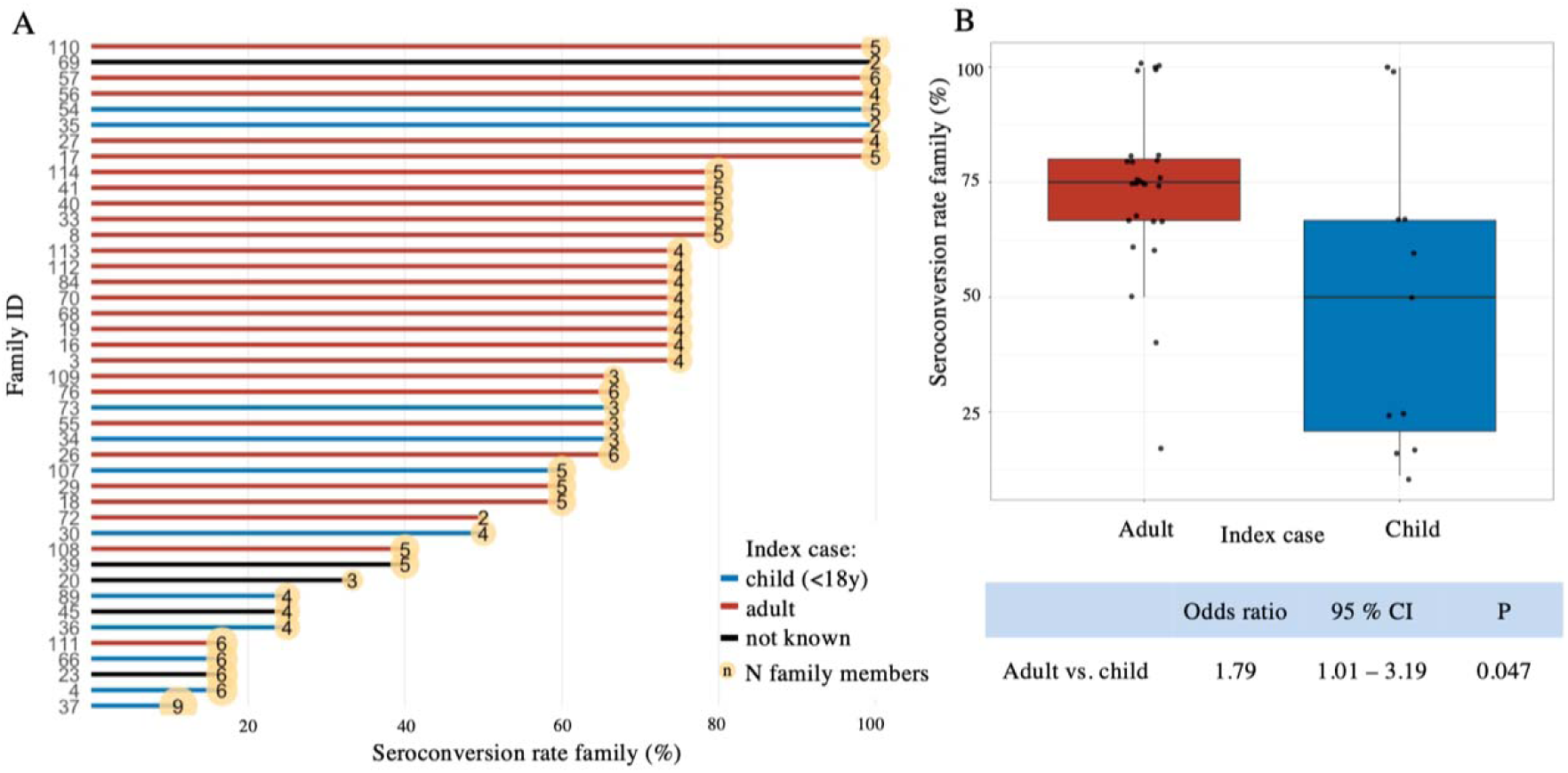
Seroconversion among household contacts. (A) Seroconversion rate of the families recalled for follow up. The seroconversion rate, calculated by the seropositive household contacts divided by all household contacts, is shown on the x-axis. Families are depicted by their respective ID and arranged by seroconversion rate from top to bottom. Families are colored according to the age group of the respective index case (child <18 years or adult >18 years), where applicable. Five families were not able to recall the index case. For two families (ID: 29, 73) at least one family member refused blood draw. (B) Seroconversion rate of families by age group of the index case. Families with an adult index case had a higher seroconversion rate compared to families with a pediatric index case (mean seroconversion rate 73·8 % vs. 48·9 %). To model the risk of seroconversion with the type of index case a logistic regression model was applied. Results are shown in the lower panel (odds ratio: 1·79, 95% CI: 1·01-3·9, P=0·047).

The follow-up phase of this study included further recall appointments for the 43 recall families, at three months and six months after screening (median 173 days, range 112-243 days and median 254 days, range 193-279 days post symptom onset, PSO). Here, PCR and serologic tests were repeated for family members. Eleven families have been lost to follow up by the end of the study, and in some cases, not all family members were present at every appointment. Four families were excluded on account of no data available on the date of symptom onset. Despite a steep rise in incidence in Hamburg, Germany in November-December 2020, no acute infection could be detected in any of the study participants by PCR.

Longitudinal analysis of antibodies showed that almost all children and adults wo were seropositive at the time of the first follow up retained detectable SARS-CoV-2 antibodies at the time of the third follow up six months later (>193 days after symptom onset). This was observed for both types of antibodies measured in this study, with nucleocapsid IgA/IgG/IgM slightly better retained in adults (seropositivity adults 100 % n=36, children 97.6 %, n=41) and spike IgG better retained in children (seropositivity adults 94.4 % n=36, children 100 %, n=41) at six months after screening in those initially positive in both tests.

The spike IgG test allowed for quantitative assessment of serum antibody concentrations. To follow changes in antibody levels of anti-spike IgG over time, statistical modelling using the zero-inflated tweedie-mixed model was applied (*Figure 2*). Time points at 90 days, 180 days and 270 days PSO were chosen for comparison between adults and children, matching the observed time frame in the study cohort (*Figure 2*). At all of these time points, children had a higher antibody concentration than adult family members (ratio children vs. adults 90 days PSO: 1.75, 95 % CI 1.40 - 2.20, P<0.001, 180 days PSO: 1.38, 95 % CI 1.08 - 1.75, P=0.01, 270 days PSO: 1.54, 95 % CI 1.19 - 1.99, P=0.001). During the observed time period, both children and adults had decreasing antibody concentrations over time. For children, the decrease was faster and more pronounced initially (*Figure 2*).

**Figure 2:**
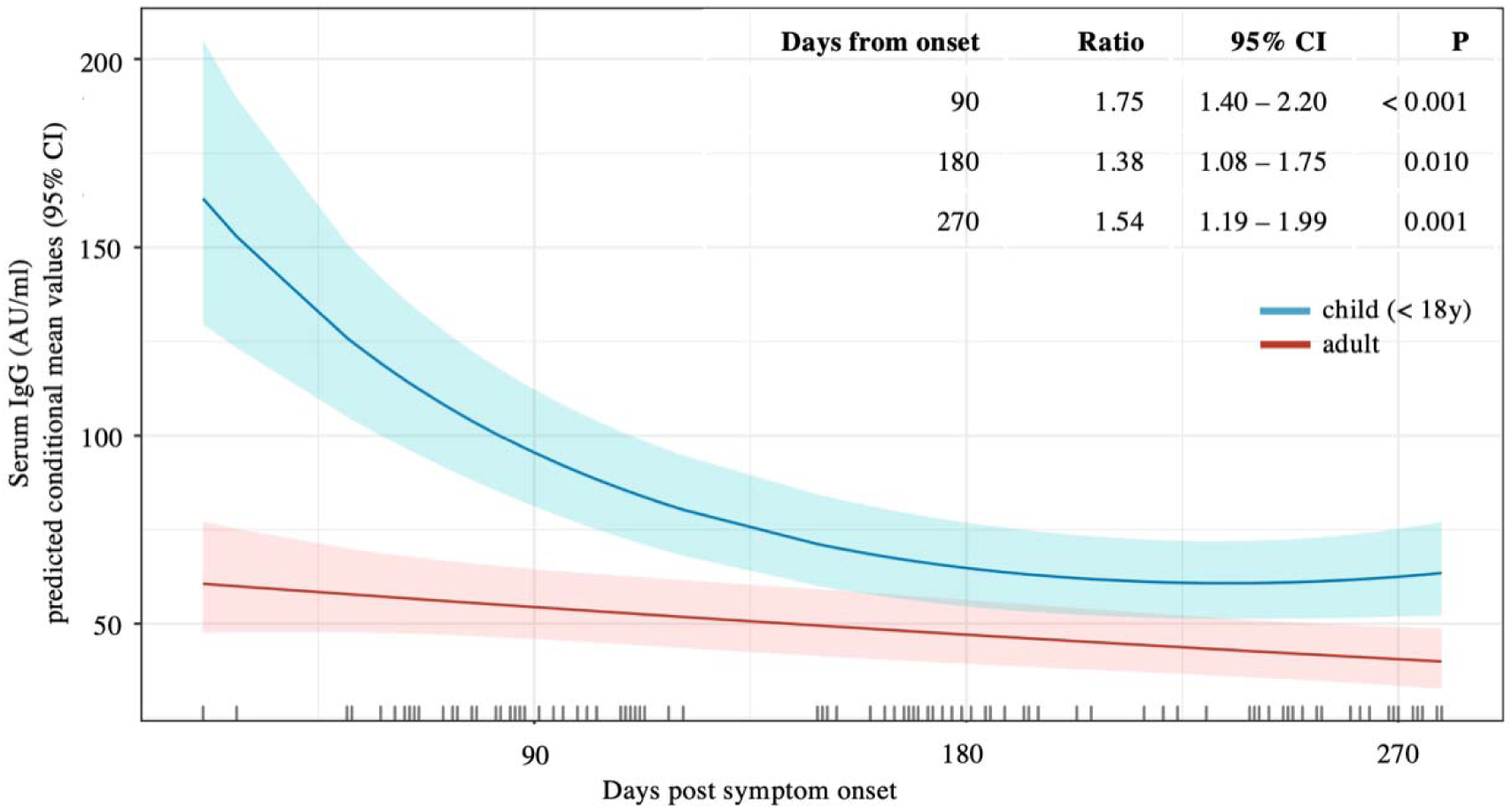
Longitudinal follow-up of serum antibody concentrations of SARS-CoV-2 anti-spike IgG in adults and children. Observation times post symptom onset are indicated on the x axis and are calculated per family. The y axis is showing predicted conditional mean values (bold curve) of the serum SARS-CoV-2 anti-spike IgG concentration as determined by the zero-inflated tweedie mixed model. The shaded areas flanking the curves indicate 95% confidence intervals. Predicted conditional means are dependent on zero-inflation. Table in the top right section showing estimated mean ratio of predicted conditional mean values children vs. adults at 90, 180 and 270 days post symptom onset, along with 95% confidence intervals and corresponding P values. Color coding for adults and children is indicated below the graph.

## Discussion

Here, we report the results of the C19.CHILD study, a large prospective seroprevalence study for SARS-CoV-2 targeting children in a low-incidence setting. Previous seroprevalence studies carried out under lockdown have compared seroconversion rates between children and adults, and found these to be lower in children [14–16]. These findings must be interpreted with caution because different contact patterns of children under lockdown may play a substantial role. A large, population-based seroepidemiological study out of Wuhan, China, a place of high initial incidence, described similar seroprevalence in children and older age groups [17]. In our cohort, seroprevalence among children increased with age, as had been the case for other cohorts of the ancestral SARS-CoV-2, including a seroprevalence study in a similar time interval from the south of Germany [14,15,18– 20]. These findings may have been specific to the ancestral variant of SARS-CoV-2 and the circumstances governing the early 2020 first wave of coronavirus infections including few imported cases followed by lockdown. Subsequent infection waves, including novel viral variants, showed an increase in seropositivity in children [21], probably based on the reopening of schools and daycare. At the same time comparison of seroprevalence in children to adults and older teenagers became less meaningful with the spread of vaccination [21].

We have found that in the case of exposure to a confirmed case of SARS-CoV-2 infection, younger children zero to under six years had a relevantly smaller risk of seroconversion than older children. Consistently, lower secondary attack rates have been reported in children than in adults [22–24]. A large study on SARS-CoV-2 transmission dynamics in two Indian states, which included children, provided evidence that contacts between people of similar age are more likely to lead to infection [25]. Thus, a lack of contact between children and their similar-age peers in a lockdown scenario might explain the differences between our data and studies in similar low-incidence settings [15,16] compared to seroprevalence studies in places with high initial incidence [17].

We report that seroconversion rates within families with index persons below the age of eighteen years were considerably lower. In our cohort, we made use of the relatively high availability of data on the index person in each family, as in the early phase of the pandemic the source of infection could still be identified in most cases. Most contact tracing and household cluster studies of SARS-CoV-2 have suggested a lower risk of transmission by children [25–30]. In a large population based cohort study spanning the whole of 2020, older children had a lower risk of transmitting SARS-CoV-2 than younger children, but a comparison with adults was not described [31]. Again, these differences may relate to the highly different contact patterns of children under a general lockdown. In Hamburg, Germany, general school and day care closure lasted from March 1 until at least May 18, 2020 and recruitment to the C19.CHILD Study began on May 11, shortly before lockdown was lifted. In this period, contacts for children outside their own families were very limited and incidence of novel SARS-CoV-2 infections was low (*Supplementary Figure S2*). Infection of most index persons in the C19.CHILD cohort probably happened before the lockdown, which is supported by patient histories regarding symptomatic illness in late February and early March (*Supplementary Table S2*). Subsequently, spread of infection would mostly have been limited to family clusters. As awareness of COVID-19 was high in the public, it may be assumed that symptomatic children, especially teenagers, would have been isolated at home (to the extent possible). The isolation of adult caregivers, however, may have been more difficult. As a result, less transmission would have occurred from children than adults. Alternatively, a less efficient biological transmission of the virus by children in the lockdown setting could explain our findings. As viral loads in nasopharyngeal swabs appear to be similar in children and adults [32,33], differences in respiratory droplet and aerosol formation [34], respiratory anatomy [35], or increased tropism of the virus in the upper vs. lower airways in children [36,37], could contribute to the suggested differences in virus transmission. It should be noted, that transmission dynamics may be different for more recent variants of SARS-CoV-2. For the B.1.617. (delta) variant, viral loads appeared to be higher than for the B.1.1.7 (alpha) variant [38], and significantly increased with age [39]. Research on the delta variant [40], and preliminary reports on the omicron variant [41,42] indicate their increased airway replication fitness compared to the ancestral and alpha variants, and this could also significantly influence the infectiousness of children with COVID-19.

In our cohort, we provide a longitudinal follow up of SARS-CoV-2 serology over nine months after symptomatic illness, showing that children as well as adults retain both anti-spike IgG and anti-nucleocapsid IgA/IgG/IgM antibodies up to 270 days post-symptom onset (PSO). To our knowledge this is the longest prospective follow-up of SARS-CoV-2 serology with a direct comparison of children and their adult household contacts reported so far. Despite the time elapsed since the beginning of the pandemic, long-term follow up of quantitative serum antibody measurements such as ours for the ancestral SARS-CoV-2 are scarce in adults and even less has been reported for children. A follow up of antibodies against SARS-CoV-2 in people living in Wuhan [17], as well as in Jiangsu [43], China, showed antibodies to persist in a majority of cases over six months. However, the authors did not provide data according to time since symptom onset or the age of the participants. Previously, persistence of antibodies in children based on two serological tests 62 days apart was reported, but time since symptom onset was also unknown [44]. One study in a somewhat comparable pediatric cohort showed also detectable IgG levels after six months and also in a single child nine months after infection [45], however the cohort did not include adult contacts for comparison. A detailed immunological follow up of adults [46] after SARS-CoV-2 infection showed similar results six to eight months PSO. Compared to adult family members, children in our cohort showed on average higher anti-spike IgG antibody levels in early reconvalescence (less than 121 days PSO). This is in contrast to earlier reports showing lower neutralizing activity [47] or similar anti-spike IgG levels [48] compared to adults, albeit addressed in older pediatric subcohorts. Pre-existing memory B cells and natural antibodies [49] or T cell memory [50,51] in children on account of more frequent common-cold coronavirus infections may lead to quicker and more efficient SARS-CoV-2 specific immune response and thus higher initial antibody concentrations. The same mechanisms might also contribute to the milder clinical course of SARS-CoV-2 that is characteristic in children. Moreover, as demonstrated in our cohort, anti-spike IgG levels remained higher in children than adults over 193 days PSO, even though a more rapid decline in serum antibody concentrations of children was observed initially.

There are limitations to this study. Teenagers were relatively underrepresented when compared to younger children and Hamburg census data. The retrospective nature of serological testing in this study did not allow for confirmation of infection by direct virus detection (PCR), therefore seropositivity is used as a surrogate marker for past infection. Furthermore, PCR test results from study participants from the time of infection were not available and index cases in family clusters were identified based on patient history of symptomatic illness (*Supplementary Table S2*). A selection bias needs to be considered when interpreting our findings. First, our study included over 50 % volunteers, second, blood draws were more likely to have been carried out in older children. In this study, no serum neutralization tests have been performed. Instead, seropositivity was defined based on two different serum antibody tests, both of which have shown good correlation with plaque reduction neutralization tests [52,53]. While this approach does not measure direct antibody mediated protective immunity, it reduces the probability of false positive tests, especially when multiple family members are assessed. Since the samples used for antibody measurements in this study were obtained after the first 2020 wave of infections, they represent infections with the ancestral SARS-CoV-2 which has now been replaced by more recent viral variants. In spite of this, our data is relevant to antibody responses after vaccination, because all current vaccines are based on inactivated virus or sequences from the ancestral SARS-CoV-2 [54–57]. The setting in which our data was obtained is unique because widespread vaccination and possible repeat infections would complicate designing similar prospective studies in the present day.

In this work we demonstrated an incomplete seroconversion of families in case of infection with the SARS-CoV-2 ancestral variant. Furthermore, seroconversion within households was lower in case of pediatric than adult index cases. Our findings showing a sustained long-term antibody responses in the pediatric population after the initial infection wave of SARS-CoV-2 is relevant for vaccination efforts targeting children as well as newly emerging viral variants.

## Data Availability

All data produced in the present study are available upon reasonable request to the authors

## Contributors

GAD, TSM, ACM, and SWG conceived the study. TSM, ACM, and SWG were responsible for study supervision. GAD, MB, MW, MKD, G Engels, KP, KF, KH, FSN, FH, LH, CK, MJK, AK, J Olfe, DEZ, ML, J Oh, TSM, ACM, SWG contributed to the study design. GAD, MB, MW, MKD, G Engels, KH, FSN, KP, FH, LJH, LH, T Kehl, CK, MJK, AK, J Olfe, F Schlenker, JS, ST, J Oh, TSM, ACM, SWG coordinated the study and acquired screening data. GAD, MW, G Engels, G Escherich, KH, MKD, KP, JB, LG, FH, LH, T Kehl, CK, MJK, RK, AK, J Olfe, F Schlenker, JS, PS, DEZ, J Oh, TSM, and ACM were responsible for recruitment. Sample preparation was supervised by GAD, MB, G Engels, KP, JB, LJH, F Sibbertsen, and LG; and GAD, MB, LJH, JS, DN, ML, and SWG were responsible for the overview of the laboratory tests. GAD, MB, MW, EV, and AZ performed statistical analysis. All authors provided conceptual input, GAD, MB, MW, ACM, and SWG wrote the essential sections of the manuscript, which was critically revised by all authors.

## References

1. Johns Hopkins University. COVID-19 Dashboard by the Center for Systems Science and Engineering (CSSE). [cited 18 Jan 2022]. Available: https://coronavirus.jhu.edu/map.html

2. Tagarro A, Epalza C, Santos M, Sanz-Santaeufemia FJ, Otheo E, Moraleda C, et al. Screening and Severity of Coronavirus Disease 2019 (COVID-19) in Children in Madrid, Spain. JAMA Pediatrics. American Medical Association; 2020. doi:10.1001/jamapediatrics.2020.1346

3. Dong Y, Mo X, Hu Y, Qi X, Jiang F, Jiang Z, et al. Epidemiological Characteristics of 2143 Pediatric Patients With 2019 Coronavirus Disease in China. Pediatrics. 2020; e20200702. doi:10.1542/peds.2020-0702

4. Götzinger F, Santiago-García B, Noguera-Julián A, Lanaspa M, Lancella L, Calò Carducci FI, et al. COVID-19 in children and adolescents in Europe: a multinational, multicentre cohort study. Lancet Child Adolesc Heal. 2020;4: 653–661. doi:10.1016/S2352-4642(20)30177-2

5. Riphagen S, Gomez X, Gonzalez-Martinez C, Wilkinson N, Theocharis P. Hyperinflammatory shock in children during COVID-19 pandemic. Lancet (London, England). 2020. doi:10.1016/S0140-6736(20)31094-1

6. Kaushik S, Aydin SI, Derespina KR, Bansal PB, Kowalsky S, Trachtman R, et al. Multisystem Inflammatory Syndrome in Children Associated with Severe Acute Respiratory Syndrome Coronavirus 2 Infection (MIS-C): A Multi-institutional Study from New York City. J Pediatr. 2020;224: 24–29. doi:10.1016/j.jpeds.2020.06.045

7. Walter EB, Talaat KR, Sabharwal C, Gurtman A, Lockhart S, Paulsen GC, et al. Evaluation of the BNT162b2 Covid-19 Vaccine in Children 5 to 11 Years of Age. N Engl J Med. 2022;386: 35–46. doi:10.1056/nejmoa2116298

8. Wilhelm A, Widera M, Grikscheit K, Toptan T, Schenk B, Pallas C, et al. Reduced Neutralization of SARS-CoV-2 Omicron Variant by Vaccine Sera and monoclonal antibodies. medRxiv. 2021; 2021.12.07.21267432. Available: https://www.medrxiv.org/content/10.1101/2021.12.07.21267432v1%0Ahttps://www.medrxiv.org/content/10.1101/2021.12.07.21267432v1.abstract

9. CDC Covid-Net. A weekly summary of U.S. COVID-19 hospitalization data; Laboratory-Confirmed COVID-19-Associated Hospitalizations. [cited 22 Jan 2022]. Available: https://gis.cdc.gov/grasp/COVIDNet/COVID19_3.html

10. Corman VM, Landt O, Kaiser M, Molenkamp R, Meijer A, Chu DK, et al. Detection of 2019 novel coronavirus (2019-nCoV) by real-time RT-PCR. Euro Surveill. 2020;25. doi:10.2807/1560-7917.ES.2020.25.3.2000045

11. Pfefferle S, Reucher S, Nörz D, Lütgehetmann M. Evaluation of a quantitative RT-PCR assay for the detection of the emerging coronavirus SARS-CoV-2 using a high throughput system. Eurosurveillance. 2020;25: 1–5. doi:10.2807/1560-7917.ES.2020.25.9.2000152

12. Centers for Disease Control and Prevention. Interim Guidelines for COVID-19 Antibody Testing. 2020. Available: https://www.cdc.gov/coronavirus/2019-ncov/lab/resources/antibody-tests-guidelines.html

13. Zhou F, Yu T, Du R, Fan G, Liu Y, Liu Z, et al. Clinical course and risk factors for mortality of adult inpatients with COVID-19 in Wuhan, China: a retrospective cohort study. Lancet. 2020. doi:10.1016/S0140-6736(20)30566-3

14. Hallal PC, Hartwig FP, Horta BL, Silveira MF, Struchiner CJ, Vidaletti LP, et al. SARS-CoV-2 antibody prevalence in Brazil: results from two successive nationwide serological household surveys. Lancet Glob Heal. 2020;8: e1390–e1398. doi:10.1016/S2214-109X(20)30387-9

15. Stringhini S, Wisniak A, Piumatti G, Azman AS, Lauer SA, Baysson H, et al. Seroprevalence of anti-SARS-CoV-2 IgG antibodies in Geneva, Switzerland (SEROCoV-POP): a population-based study. Lancet. 2020;396: 313–319. doi:10.1016/S0140-6736(20)31304-0

16. Pollán M, Pérez-Gómez B, Pastor-Barriuso R, Oteo J, Hernán MA, Pérez-Olmeda M, et al. Prevalence of SARS-CoV-2 in Spain (ENE-COVID): a nationwide, population-based seroepidemiological study. Lancet. 2020;396: 535–544. doi:10.1016/S0140-6736(20)31483-5

17. He Z, Ren L, Yang J, Guo L, Feng L, Ma C, et al. Seroprevalence and humoral immune durability of anti-SARS-CoV-2 antibodies in Wuhan, China: a longitudinal, population-level, cross-sectional study. Lancet. 2021;397: 1075–1084. doi:10.1016/S0140-6736(21)00238-5

18. Reicher S, Ratzon R, Ben-Sahar S, Hermoni-Alon S, Mossinson D, Shenhar Y, et al. Nationwide seroprevalence of antibodies against SARS-CoV-2 in Israel. Eur J Epidemiol. 2021. doi:10.1007/s10654-021-00749-1

19. Tönshoff B, Müller B, Elling R, Renk H, Meissner P, Hengel H, et al. Prevalence of SARS-CoV-2 Infection in Children and Their Parents in Southwest Germany. JAMA Pediatr. 2021. doi:10.1001/jamapediatrics.2021.0001

20. Viner RM, Mytton OT, Bonell C, Melendez-Torres GJ, Ward J, Hudson L, et al. Susceptibility to SARS-CoV-2 Infection among Children and Adolescents Compared with Adults: A Systematic Review and Meta-analysis. JAMA Pediatrics. American Medical Association; 2020. doi:10.1001/jamapediatrics.2020.4573

21. Oeser C, Whitaker H, Linley E, Borrow R, Tonge S, Brown CS, et al. Large increases in SARS-CoV-2 seropositivity in children in England: Effects of the delta wave and vaccination. J Infect. 2021; 19–21. doi:10.1016/j.jinf.2021.11.019

22. Yung CF, Kam K qian, Chong CY, Nadua KD, Li J, Tan NWH, et al. Household Transmission of Severe Acute Respiratory Syndrome Coronavirus 2 from Adults to Children. J Pediatr. 2020;225: 249–251. doi:10.1016/j.jpeds.2020.07.009

23. Madewell ZJ, Yang Y, Longini IM, Halloran ME, Dean NE. Household Transmission of SARS-CoV-2: A Systematic Review and Meta-analysis. JAMA Netw open. 2020;3: e2031756. doi:10.1001/jamanetworkopen.2020.31756

24. Fung HF, Martinez L, Alarid-Escudero F, Salomon JA, Studdert DM, Andrews JR, et al. The household secondary attack rate of severe acute respiratory syndrome coronavirus 2 (sars-cov-2): A rapid review. Clin Infect Dis. 2021;73: S138–S145. doi:10.1093/cid/ciaa1558

25. Laxminarayan R, Wahl B, Dudala SR, Gopal K, Mohan C, Neelima S, et al. Epidemiology and transmission dynamics of COVID-19 in two Indian states. Science (80-). 2020; eabd7672. doi:10.1126/science.abd7672

26. Merckx J, Labrecque JA, Kaufman JS. Transmission of SARS-CoV-2 by Children. Dtsch Arztebl Int. 2020;117: 553–560. Available: https://www.aerzteblatt.de/int/article.asp?id=214818

27. Yi S, Kim YM, Choe YJ, Ahn S, Han S, Park YJ. Geospatial Analysis of Age-specific SARS-CoV-2 Transmission Patterns in Households, Korea. J Korean Med Sci. 2021;36: 1–3. doi:10.3346/jkms.2021.36.e63

28. Grijalva CG, Rolfes MA, Zhu Y, Mclean HQ, Hanson KE, Belongia; Edward A, et al. Morbidity and Mortality Weekly Report Transmission of SARS-COV-2 Infections in Households-Tennessee and Wisconsin, April-September 2020. 2020. Available: https://www.cdc.gov/mmwr/volumes/69/wr/pdfs/mm6944e1-H.pdf

29. Soriano-Arandes A, Gatell A, Serrano P, Biosca M, Campillo F, Capdevila R, et al. Household Severe Acute Respiratory Syndrome Coronavirus 2 Transmission and Children: A Network Prospective Study. Clin Infect Dis. 2021;73: e1261–e1269. doi:10.1093/cid/ciab228

30. Brandal LT, Ofitserova TS, Meijerink H, Rykkvin R, Lund HM, Hungnes O, et al. Minimal transmission of SARS-CoV-2 from paediatric COVID-19 cases in primary schools, Norway, August to November 2020. Eurosurveillance. 2020;26: 3–8. doi:10.2807/1560-7917.ES.2020.26.1.2002011

31. Paul LA, Daneman N, Schwartz KL, Science M, Brown KA, Whelan M, et al. Association of Age and Pediatric Household Transmission of SARS-CoV-2 Infection. JAMA Pediatr. 2021;175: 1151–1158. doi:10.1001/jamapediatrics.2021.2770

32. Jones TC, Biele G, Mühlemann B, Veith T, Schneider J, Beheim-Schwarzbach J, et al. Estimating infectiousness throughout SARS-CoV-2 infection course. Science (80-). 2021;373. doi:10.1126/science.abi5273

33. Baggio S, L’Huillier AG, Yerly S, Bellon M, Wagner N, Rohr M, et al. Severe Acute Respiratory Syndrome Coronavirus 2 (SARS-CoV-2) Viral Load in the Upper Respiratory Tract of Children and Adults with Early Acute Coronavirus Disease 2019 (COVID-19). Clin Infect Dis. 2021;73: 148–150. doi:10.1093/cid/ciaa1157

34. Yang S, Lee GWM, Chen CM, Wu CC, Yu KP. The size and concentration of droplets generated by coughing in human subjects. J Aerosol Med Depos Clear Eff Lung. 2007;20: 484–494. doi:10.1089/jam.2007.0610

35. Wald ER, Schmit KM, Gusland DY. A Pediatric Infectious Disease Perspective on COVID-19. Clin Infect Dis. 2020. doi:10.1093/cid/ciaa1095

36. Saheb Sharif-Askari N, Saheb Sharif-Askari F, Alabed M, Temsah MH, Al Heialy S, Hamid Q, et al. Airways Expression of SARS-CoV-2 Receptor, ACE2, and TMPRSS2 Is Lower in Children Than Adults and Increases with Smoking and COPD. Mol Ther - Methods Clin Dev. 2020;18: 1–6. doi:10.1016/j.omtm.2020.05.013

37. Muus C, Luecken MD, Eraslan G, Waghray A, Heimberg G, Sikkema L, et al. Integrated analyses of single-cell atlases reveal age, gender, and smoking status associations with cell type-specific expression of mediators of SARS-CoV-2 viral entry and highlights inflammatory programs in putative target cells. bioRxiv. 2020; 2020.04.19.049254. doi:10.1101/2020.04.19.049254

38. Costa R, Olea B, Bracho MA, Albert E, de Michelena P, Martínez-Costa C, et al. RNA viral loads of SARS-CoV-2 Alpha and Delta variants in nasopharyngeal specimens at diagnosis stratified by age, clinical presentation and vaccination status. J Infect. 2021; 29–31. doi:10.1016/j.jinf.2021.12.018

39. Singanayagam A, Hakki S, Dunning J, Madon KJ, Crone MA, Koycheva A, et al. Community transmission and viral load kinetics of the SARS-CoV-2 delta (B.1.617.2) variant in vaccinated and unvaccinated individuals in the UK: a prospective, longitudinal, cohort study. Lancet Infect Dis. 2021;3099. doi:10.1016/S1473-3099(21)00648-4

40. Mlcochova P, Kemp S, Dhar MS, Papa G, Meng B, Ferreira IATM, et al. SARS-CoV-2 B.1.617.2 Delta variant replication and immune evasion. Nature. 2021;599. doi:10.1038/s41586-021-03944-y

41. Chan MCW, Hui KPY, Ho J, Cheung M, Ng K, Ching R, et al. SARS-CoV-2 Omicron variant replication in human respiratory tract ex vivo. Nat Portf. 2022. doi:10.21203/rs.3.rs-1189219/v1

42. Peacock TP, Brown JC, Zhou J, Thakur N, Newman J, Kugathasan R, et al. The SARS-CoV-2 variant, Omicron, shows rapid replication in human primary nasal epithelial cultures and efficiently uses the endosomal route of entry. bioRxiv. 2022; 2021.12.31.474653. doi:10.1101/2021.12.31.474653

43. Zhu L, Xu X, Zhu B, Guo X, Xu K, Song C, et al. Kinetics of SARS-CoV-2 Specific and Neutralizing Antibodies over Seven Months after Symptom Onset in COVID-19 Patients. Microbiol Spectr. 2021;9. doi:10.1128/spectrum.00590-21

44. Roarty C, Tonry C, McFetridge L, Mitchell H, Watson C, Waterfield T, et al. Kinetics and seroprevalence of SARS-CoV-2 antibodies in children. The Lancet Infectious Diseases. Lancet Publishing Group; 2020. doi:10.1016/S1473-3099(20)30884-7

45. Oygar PD, Ozsurekci Y, Gurlevik SL, Aykac K, Kukul MG, Cura Yayla BC, et al. Longitudinal Follow-up of Antibody Responses in Pediatric Patients with COVID-19 up to 9 Months after Infection. Pediatr Infect Dis J. 2021;40: E294–E299. doi:10.1097/INF.0000000000003199

46. Dan JM, Mateus J, Kato Y, Hastie KM, Yu ED, Faliti CE, et al. Immunological memory to SARS-CoV-2 assessed for up to 8 months after infection. Science (80-). 2021;371. doi:10.1126/science.abf4063

47. Pierce CA, Preston-Hurlburt P, Dai Y, Burn Aschner C, Cheshenko N, Galen B, et al. Immune responses to SARS-CoV-2 infection in hospitalized pediatric and adult patients. Sci Transl Med. 2020. Available: http://stm.sciencemag.org/

48. Weisberg SP, Connors TJ, Zhu Y, Baldwin MR, Lin W-H, Wontakal S, et al. Distinct antibody responses to SARS-CoV-2 in children and adults across the COVID-19 clinical spectrum. Nat Immunol. 2020. doi:10.1038/s41590-020-00826-9

49. Carsetti R, Quintarelli C, Quinti I, Piano Mortari E, Zumla A, Ippolito G, et al. The immune system of children: the key to understanding SARS-CoV-2 susceptibility? The Lancet Child and Adolescent Health. Elsevier B.V.; 2020. pp. 414–416. doi:10.1016/S2352-4642(20)30135-8

50. Sette A, Crotty S. Pre-existing immunity to SARS-CoV-2: the knowns and unknowns. Nature Reviews Immunology. Nature Research; 2020. pp. 457–458. doi:10.1038/s41577-020-0389-z

51. Braun J, Loyal L, Frentsch M, Wendisch D, Georg P, Kurth F, et al. SARS-CoV-2-reactive T cells in healthy donors and patients with COVID-19. Nature. 2020;587: 270–274. doi:10.1038/s41586-020-2598-9

52. Kohmer N, Westhaus S, Rühl C, Ciesek S, Rabenau HF. Brief clinical evaluation of six high-throughput SARS-CoV-2 IgG antibody assays. J Clin Virol. 2020;129. doi:10.1016/j.jcv.2020.104480

53. Bahar B, Jacquot C, Mo YD, DeBiasi RL, Campos J, Delaney M. Kinetics of Viral Clearance and Antibody Production Across Age Groups in Children with Severe Acute Respiratory Syndrome Coronavirus 2 Infection. J Pediatr. 2020;227: 31–37.e1. doi:10.1016/j.jpeds.2020.08.078

54. Mulligan MJ, Lyke KE, Kitchin N, Absalon J, Gurtman A, Lockhart S, et al. Phase I/II study of COVID-19 RNA vaccine BNT162b1 in adults. Nature. 2020;586: 589–593. doi:10.1038/s41586-020-2639-4

55. Corbett KS, Edwards DK, Leist SR, Abiona OM, Boyoglu-Barnum S, Gillespie RA, et al. SARS-CoV-2 mRNA vaccine design enabled by prototype pathogen preparedness. Nature. 2020;586: 567–571. doi:10.1038/s41586-020-2622-0

56. Xia S, Zhang Y, Wang Y, Wang H, Yang Y, Gao GF, et al. Safety and immunogenicity of an inactivated SARS-CoV-2 vaccine, BBIBP-CorV: a randomised, double-blind, placebo-controlled, phase 1/2 trial. Lancet Infect Dis. 2021;21: 39–51. doi:10.1016/S1473-3099(20)30831-8

57. Wang H, Zhang Y, Huang B, Deng W, Quan Y, Wang W, et al. Development of an Inactivated Vaccine Candidate, BBIBP-CorV, with Potent Protection against SARS-CoV-2. Cell. 2020;182: 713–721.e9. doi:10.1016/j.cell.2020.06.008

