## Supplementary Material for "Long-term antibody response to SARS-CoV-2 in children"

**Hamburg, Germany**

University Medical Center Hamburg-Eppendorf, Department of Pediatrics: Lea Bandel, Markus Baumanns, Jan Beime, M.D., Pia Dähler, Gabor A. Dunay, M.D., Ph.D., Barbara Dwenger, Geraldine Engels, M.D., Ph.D., Annika Erdmann, Lev Grinstein, M.D., Laura Hecher, M.D., Sophia Hegselmann, Katharina Hermann, M.D., Kai Hornig, Johanna Jipp, Pia Kirkerup, Aloisa Kohl, M.D., Michael Krumm, Pelin Kurnaz M.D., Ania C. Muntau, M.D., Friederike S. Neumann, M.D., Jun Oh, M.D., Sabine Pasterkamp, Kevin Paul, M.D., Friderike Schlenker, M.D., Anna Serve, Daniel Tegtmeyer, M.D., Julia Terstegen, Ceri Theresa Wiedling, M.D., Madelaine Wingerath, Mathias Woidy, M.D., Dimitra E. Zazara, M.D. Ph.D.

University Medical Center Hamburg-Eppendorf, University Children’s Research: Madalena Barroso, Ph.D., Marta K. Danecka, Ph.D., Gabor A. Dunay, M.D., Ph.D., Geraldine Engels, M.D., Ph.D., Stefanie Etzold, Søren W. Gersting, M.D., Ingrid Goebel, Armin Günther, Luka J. Haupt, Pia-Sophie Kantor, Thomas Klokow, Ph.D., Friederike S. Neumann, M.D., Kevin Paul, M.D., Jessica Schmiesing, Ph.D., Freya Sibbersten, Stephan Tiede, Ph.D., Kristin Fehse, Mathias Woidy, M.D.

University Medical Center Hamburg-Eppendorf, Department of Pediatric Cardiology: Florian Arndt, M.D., Stefan Blankenberg, M.D., Daniel Diaz, M.D., Franziska Haniel, M.D., Torben Kehl, M.D., Peer Hauck, M.D., Rainer G. Kozlik-Feldmann, M.D., Thomas S. Mir, M.D., Götz C. Müller, M.D., Jakob Olfe, M.D., Veronika C. Stark, M.D., Peter Wiegand

University Medical Center Hamburg-Eppendorf, Pediatric Hematology and Oncology: Gabriele Escherich, M.D., Stefan Rutkowski, M.D., Johanna Schrum M.D., Beate Winkler, M.D.

University Medical Center Hamburg-Eppendorf, Department of Pediatric Stem Cell Transplantation: Bernd Hartz, M.D., Anne Kruchen, Ph.D., Ingo Müller, M.D.

University Medical Center Hamburg-Eppendorf, Department of Pediatric Surgery: Michael Boettcher, M.D., Konrad Reinshagen, M.D., Carolin Stiel, M.D., Katharina Wenke, M.D.

University Medical Center Hamburg-Eppendorf, Neonatology and Intensive Care Unit: Joana Adler Fernandes de Abreu, M.D., Marlies Bergers, Martin Blohm, M.D., Philipp Deindl, M.D., Theresa Harbauer, M.D., Cornelius Rau, M.D., Dominique Singer, M.D.

University Medical Center Hamburg-Eppendorf, Department of Obstetrics and Fetal Medicine: Petra Arck, M.D., Anke Diemert, M.D., Corinna Cramer, M.D., Kurt Hecher, M.D.

University Medical Center Hamburg-Eppendorf, Department of Pediatric Hematology and Oncology. Research Institute Children's Cancer Center: Marianne Klokow, Julia Strauss

Altona Children’s Hospital: Anja Große Lordemann, M.D., Maria-Dorothee Neumann, Philippe Stock, M.D.

Wilhelmstift Children’s Hospital: Peter Höger, M.D., Christoph Kemen, M.D.

Helios Mariahilf Klinik: Caroline Schmitt, M.D.

Asklepios Klinik Nord-Heidberg, Klinik und Poliklinik für Kinder- und Jugendmedizin: Markus J. Kemper, M.D.

University Medical Center Hamburg-Eppendorf, University Heart and Vascular Center, Epidemiologic Study Center: Stefan Blankenberg, M.D., Ines Schäfer, Ph.D.

University Medical Center Hamburg-Eppendorf, Business Division for Information Technology: Jens Vogel, Maximilian Noelle-Wying

University Medical Center Hamburg-Eppendorf, Institute of Medical Microbiology, Virology and Hygiene: Martin Aepfelbacher, M.D., Kathrin Cermann, Armin Hoffmann, M.D., Johannes K.-M. Knobloch, M.D., Marc Lütgehetmann, M.D., Dominik Nörz, M.D.

University Medical Center Hamburg-Eppendorf, First Department of Medicine, Division of Infectious Diseases: Marylyn M. Addo, M.D., Robin Kobbe, M.D., Julian Schulze zur Wiesch, M.D.

University Medical Center Hamburg-Eppendorf, Institute of Medical Biometry and Epidemiology: Eik Vettorazzi, Antonia Zapf, M.D.

University Medical Center Hamburg-Eppendorf, Institute of Human Genetics: Davor Lessel, M.D., Ph.D.

Berlin, Germany

Institute of Virology, Charité Universitätsmedizin Berlin, Berlin, Germany: Christian Drosten, M.D.

### **Supplementary Methods**

### **Study cohort**

The C19.CHILD study was conducted in accordance with the guidelines of the Declaration of Helsinki and approved by the Ethical Committee of the Hamburg Chamber of Physicians (ethical review number PV7336).

The study population consisted of i) in- and outpatients aged 0 to <18 years of all pediatric hospitals in Hamburg, Germany ii) healthy children aged 0 to <18 years as volunteers iii) participants of other studies (Prenatal Investigation of Children’s Health- PRINCE study, Hamburg City Health Study) aged 0 to <18 years, iv) children, who were household contacts of SARS-CoV-2 positive study participants.

A written informed consent was obtained from parents or guardians in all cases, from children over 7 years whenever possible, for children also consent in spoken word was accepted.

The inclusion criteria were i) children or teenagers aged 0-18 years ii) patient in one of the participating centers or volunteer in the central C19.CHILD Study Clinic iii) informed consent from parents or guardians iv) informed consent from children >7 years (unless not capable)

The exclusion criteria were i) prematurity <37 weeks of gestation ii) informed consent of parents or guardians not possible in spoken word or otherwise iii) informed consent not given.

#### **Clinical samples**

Nasopharyngeal swabs were collected and preserved in 3 ml of universal transport medium (UTM; Miraclean Technology). All swab samples were inactivated and used for viral RNA isolation within 72 hours. Blood samples were processed within 24 hours for serum separation.

**Viral RNA extraction and qPCR**

RNA was extracted from clinical samples using the NucleoSpin 96 Virus Core Kit (Macherey-Nagel). Samples were inactivated upon addition of the kit lysis buffer (RAV-1) in 1:4 ratio (v/v) and incubated for 30 min before RNA isolation. The kit manufacturer´s instructions were adapted for automated RNA extraction using a Tecan Freedom Evo liquid handler system equipped with a TeVacs vacuum unit (Tecan).

E- and S-gene quantitative PCR was used for qualitative detection of lineage B-betacoronavirus (B-βCoV) and SARS-CoV-2. RNA reverse transcription and PCR were performed using RealStar SARS-CoV-2 RT-PCR Kit 1.0 (altona Diagnostics). An RNA internal control (RealStar kit component) was added to the inactivated samples prior to RNA isolation for control of RNA isolation and RT-PCR inhibition. Multiplex qPCR using distinct probe dyes for the viral E-gene (B-βCoV specific), the S-gene (SARS-CoV-2 specific), and the internal control, allowed for parallel detection of the different targets. The RT-PCR reaction comprised a 30 µL reaction containing 10 µL of extracted RNA and 20 µL of RT-PCR reaction mix. Thermal cycling consisted of an incubation at 55 °C for 10 min for reverse transcription, followed by 95 °C for 2 min and then 45 cycles of 95 °C for 15 sec, 55 °C for 45 sec and 72 °C for 15 sec. A QuantStudio 12K Flex Real-Time PCR System equipped with a standard 96-well block (Applied Biosystems) or a CFX96 Connect cycler (Bio-Rad Laboratories) were used. *In vitro* transcribed E-gene RNA was spiked into RAV-1 before RNA isolation and used as a positive control, while the negative control consisted of UTM in RAV-1. In addition, an E- and S-gene positive control provided by the RealStar kit was used in the RT-PCR reaction.

The assessment of the limit of detection (LOD) of the test was done using 10 replicates of a dilution series of EDX SARS-CoV-2 synthetic RNA (Exact Diagnostics) containing five gene targets (E, N, ORF1ab, RdRP and S Genes of SARS-CoV-2) in UTM and RAV-1. The LOD was 156 copies/mL for SARS-CoV-2 specific S-gene, and 313 copies/mL for B-ßCoV E-gene, with a detection rate over 95 %.

For RT-PCR data analysis, QuantStudio 12K Flex (Applied Biosystems) or CFX Manager (Bio-Rad Laboratories) software were used. Samples were considered valid if the Ct value for the internal control was lower than 35. Amplification of S-/E-gene, independent of the Ct, was considered positive.

#### **Serology tests**

After serum separation, samples were directly used for antibody testing or stored at −20 °C prior to analysis.

Sample screening was performed using the Elecsys® AntiSARS-CoV-2 assay (Roche) in a cobas e411 system and a cut-off index above 1 was used to define positivity, following the manufacturer’s instructions. Elecsys® AntiSARS-CoV-2 detects antibodies (IgA, IgM, IgG) against the SARS-CoV-2 nucleocapsid protein with a 99.8 % specificity (according to Roche).

The LIAISON® SARS-CoV-2 S1/S2 IgG assay (DiaSorin) was used as a confirmatory assay. LIAISON® assay was performed using a Liaison XL system, following the manufacturers’ recommendations. 10 AU/mL was used as the cut-off for positivity. Assay specificity is 98.5 %, according to the manufacturer, for detection of IgG antibodies against the spike S1 and S2 proteins.

### **Supplementary Tables and Figures**

**Table S1. Underlying medical conditions in C19.CHILD study population**

Distribution of participants with underlying medical conditions in the population is presented as counts (percent). Data is compared using Fisher’s exact test.

|  | **Negative in either test (N=4590)** | **Positive Roche & DiaSorin (N=67)** | **Total  (N=4657)** | **P value** |
| --- | --- | --- | --- | --- |
| Underlying condition | 1334 (29.1 % %) | 16 (23.9 %) | 1350 (29.0 %) | 0.42 |
| thereof |  |  |  |  |
| Respiratory | 157 (11.8 %) | 2 (12.5 %) | 159 (11.8 %) | >0.99 |
| Hepatic | 42 (3.1 %) | 0 (0.0 %) | 42 (3.1 %) | >0.99 |
| Onco-/Hematologic | 220 (16.5 %) | 1 (6.3 %) | 221 (16.4 %) | 0.38 |
| Immunologic | 91 (6.8 %) | 2 (12.5 %) | 93 (6.9 %) | 0.39 |
| Neurologic | 163 (12.2 %) | 1 (6.3 %) | 164 (12.1 %) | 0.73 |
| Rheumatologic | 63 (4.7 %) | 1 (6.3 %) | 64 (4.7 %) | 0.61 |
| Cardiovascular | 216 (16.2 %) | 3 (18.8 %) | 219 (16.2 %) | >0.99 |
| Metabolic | 122 (9.1 %) | 1 (6.3 %) | 123 (9.1 %) | >0.99 |
| Renal | 164 (12.3 %) | 1 (6.3 %) | 165 (12.2 %) | 0.73 |
| Inflammatory Bowel Disease | 53 (4.0 %) | 1 (6.3 %) | 54 (4.0 %) | 0.54 |
| Endocrine | 32 (2.4 %) | 0 (0.0 %) | 32 (2.4 %) | >0.99 |
| Atopic | 106 (7.9 %) | 3 (18.8 %) | 109 (8.1 %) | 0.21 |
| Liver Transplantation | 21 (1.6 %) | 0 (0.0 %) | 21 (1.6 %) | >0.99 |
| Kidney Transplantation | 24 (1.8 %) | 0 (0.0 %) | 24 (1.8 %) | >0.99 |
| Transplantation | 80 (6.0 %) | 0 (0.0 %) | 80 (5.9 %) | 0.63 |
| Hematopoietic Stem Cell Transplantation | 24 (1.8 %) | 0 (0.0 %) | 24 (1.8 %) | >0.99 |
| Solid Organ Transplantation | 46 (3.4 %) | 0 (0.0 %) | 46 (3.4 %) | >0.99 |
| Psychologic | 18 (1.3 %) | 0 (0.0 %) | 18 (1.3 %) | >0.99 |
| Trisomy 21 | 11 (0.8 %) | 0 (0.0 %) | 11 (0.8 %) | >0.99 |

#### **Table S2. Overview of the 43 families recalled for the follow-up phase.**

Three children refused repeated blood draw in the follow-up phase. However, for the calculation of the families’ seroconversion rate the result from the screening phase was taken (participants are marked with*). For two families one family member refused blood draw (Family IDs: 29,73).

| **Family ID** | **DiaSorin**  **IgG** | **Roche**  **IgM** | **Age (y)** | **Relationship** | **Self-reported symptoms** | **Time of symptomatic illness** | **Index case** |
| --- | --- | --- | --- | --- | --- | --- | --- |
| 3 | positive | positive | 2 | screening positive | yes |  | parent |
| 3* | negative | negative | 6 | sibling | no |  |  |
| 3 | positive | positive | 38 | parent | NA |  |  |
| 3 | positive | positive | 38 | parent | yes | March 2020 |  |
| 4 | positive | positive | 17 | screening positive | yes | March 2020 | screening positive |
| 4 | negative | negative | 14 | sibling | no |  |  |
| 4 | negative | negative | 12 | sibling | no |  |  |
| 4 | negative | negative | 57 | parent | no |  |  |
| 4 | negative | negative | 19 | sibling | no |  |  |
| 4 | negative | negative | 52 | parent | no |  |  |
| 8 | positive | positive | 6 | screening positive | no |  | parent |
| 8 | positive | positive | 10 | screening positive | no |  |  |
| 8 | positive | positive | 44 | parent | yes |  |  |
| 8 | negative | negative | 78 | grandparent | yes |  |  |
| 8 | positive | positive | 42 | parent | yes |  |  |
| 16 | positive | positive | 1 | screening positive | yes | March 2020 | parent |
| 16 | negative | negative | 3 | sibling | yes |  |  |
| 16 | positive | positive | 34 | parent | yes | March 2020 |  |
| 16 | positive | positive | 33 | parent | yes | March 2020 |  |
| 17 | positive | positive | 15 | screening positive | no |  | sibling (>18y) |
| 17 | positive | positive | 55 | parent | no |  |  |
| 17 | positive | positive | 49 | parent | no |  |  |
| 17 | positive | positive | 23 | sibling | no |  |  |
| 17 | positive | positive | 19 | sibling | yes | March 2020 |  |
| 18 | positive | positive | 7 | screening positive | yes | March 2020 | parent |
| 18 | negative | positive | 3 | screening positive | no |  |  |
| 18 | negative | negative | 9 | sibling | no |  |  |
| 18 | positive | positive | 49 | parent | yes | March 2020 |  |
| 18 | positive | positive | 39 | parent | yes | March 2020 |  |
| 19 | positive | positive | 6 | screening positive | no |  | parent |
| 19 | positive | positive | 11 | screening positive | no |  |  |
| 19 | positive | positive | 47 | parent | yes |  |  |
| 19 | negative | negative | 46 | parent | yes | March 2020 |  |
| 20 | positive | positive | 15 | screening positive | no |  | NA |
| 20 | negative | negative | 63 | parent | no |  |  |
| 20 | negative | negative | 56 | parent | no |  |  |
| 23 | positive | positive | 13 | screening positive | yes | March 2020 | NA |
| 23 | negative | negative | 4 | sibling | yes |  |  |
| 23 | negative | negative | 1 | sibling | yes |  |  |
| 23 | negative | negative | 9 | sibling | yes |  |  |
| 23 | negative | negative | 39 | parent | yes |  |  |
| 23 | negative | negative | 35 | parent | yes |  |  |
| 26 | positive | positive | 7 | screening positive | no |  | parent |
| 26 | negative | negative | 3 | sibling | no |  |  |
| 26 | positive | positive | 3 | sibling | no |  |  |
| 26 | negative | negative | 47 | parent | no |  |  |
| 26 | positive | positive | 53 | parent | yes | March 2020 |  |
| 26 | positive | positive | 24 | other | no |  |  |
| 27 | positive | positive | 0 | screening positive | no |  | parent |
| 27 | positive | positive | 6 | sibling | no |  |  |
| 27 | positive | positive | 35 | parent | yes | March 2020 |  |
| 27 | positive | positive | 32 | parent | yes | March 2020 |  |
| 29 | positive | positive | 10 | screening positive | no |  | parent |
| 29 |  |  | 3 | sibling | no |  |  |
| 29 | negative | negative | 7 | sibling | no |  |  |
| 29 | positive | positive | 42 | parent | yes | April 2020 |  |
| 29 | positive | positive | 45 | parent | yes | April 2020 |  |
| 30 | positive | positive | 12 | screening positive | no |  | sibling |
| 30 | negative | negative | 11 | sibling | yes | March 2020 |  |
| 30 | positive | positive | 44 | parent | yes | March 2020 |  |
| 30 | negative | positive | 45 | parent | NA |  |  |
| 33 | positive | positive | 16 | screening positive | yes | April 2020 | parent |
| 33 | positive | positive | 11 | screening positive | yes | April 2020 |  |
| 33 | positive | positive | 14 | screening positive | yes |  |  |
| 33 | positive | positive | 50 | parent | yes | April 2020 |  |
| 33 | negative | positive | 50 | parent | yes | March 2020 |  |
| 34 | positive | positive | 11 | screening positive | no |  | screening positive |
| 34 | positive | positive | 48 | parent | no |  |  |
| 34 | negative | negative | 46 | parent | no |  |  |
| 35 | positive | positive | 3 | screening positive | yes | March 2020 | screening positive |
| 35 | positive | positive | 39 | parent | yes | March 2020 |  |
| 36 | positive | positive | 17 | screening positive | yes | March 2020 | screening positive |
| 36 | negative | negative | 12 | sibling | no |  |  |
| 36 | negative | negative | 50 | parent | no |  |  |
| 36 | negative | negative | 51 | parent | no |  |  |
| 37 | positive | positive | 10 | screening positive | yes | March 2020 | screening positive |
| 37 | positive | negative | 3 | sibling | yes | March 2020 |  |
| 37 | negative | negative | 4 | sibling | yes | March 2020 |  |
| 37 | negative | negative | 29 | other | yes | March 2020 |  |
| 37 | negative | negative | 55 | parent | yes | March 2020 |  |
| 37 | negative | negative | 30 | sibling | yes | March 2020 |  |
| 37 | negative | negative | 76 | grandparent | no |  |  |
| 37 | negative | negative | 53 | parent | no |  |  |
| 37 | negative | negative | 19 | sibling | yes | March 2020 |  |
| 39 | positive | positive | 15 | screening positive | yes |  | NA |
| 39 | negative | negative | 12 | sibling | no |  |  |
| 39 | positive | positive | 9 | sibling | no |  |  |
| 39 | negative | negative | 44 | parent | yes |  |  |
| 39 | negative | negative | 45 | parent | yes |  |  |
| 40 | positive | positive | 7 | screening positive | yes |  | parent |
| 40 | positive | positive | 14 | screening positive | yes | March 2020 |  |
| 40 | negative | negative | 11 | sibling | yes |  |  |
| 40 | positive | positive | 44 | parent | yes | March 2020 |  |
| 40 | positive | positive | 44 | parent | yes | March 2020 |  |
| 41 | positive | positive | 11 | screening positive | no |  | parent |
| 41 | positive | positive | 20 | sibling | yes | March 2020 |  |
| 41 | negative | negative | 23 | sibling | yes | March 2020 |  |
| 41 | positive | positive | 53 | parent | yes | March 2020 |  |
| 41 | positive | positive | 51 | parent | yes |  |  |
| 45 | positive | positive | 9 | screening positive | no |  | NA |
| 45 | negative | positive | 13 | screening positive | no |  |  |
| 45 | negative | negative | 53 | parent | no |  |  |
| 45 | negative | negative | 47 | parent | no |  |  |
| 54 | positive | positive | 13 | screening positive | no |  | screening positive |
| 54* | positive | positive | 13 | screening positive | yes |  |  |
| 54 | positive | positive | 16 | screening positive | yes | March 2020 |  |
| 54 | positive | positive | 49 | parent | yes | March 2020 |  |
| 54 | positive | positive | 52 | parent | no |  |  |
| 55 | positive | positive | 6 | screening positive | yes | April 2020 | parent |
| 55 | negative | negative | 44 | parent | yes | March 2020 |  |
| 55 | positive | positive | 41 | parent | no |  |  |
| 56 | positive | positive | 12 | screening positive | no |  | parent |
| 56 | positive | positive | 5 | sibling | yes | March 2020 |  |
| 56 | positive | positive | 47 | parent | yes | March 2020 |  |
| 56 | positive | positive | 46 | parent | yes | March 2020 |  |
| 57 | positive | positive | 15 | screening positive | no |  | parent |
| 57 | positive | positive | 17 | sibling | no |  |  |
| 57 | positive | positive | 10 | sibling | no |  |  |
| 57 | positive | positive | 3 | sibling | no |  |  |
| 57 | positive | positive | 52 | parent | yes | May 2020 |  |
| 57 | positive | positive | 43 | parent | yes | May 2020 |  |
| 66 | positive | positive | 17 | screening positive | yes | March 2020 | sibling |
| 66 | negative | negative | 12 | sibling | yes | March 2020 |  |
| 66 | negative | negative | 12 | sibling | no |  |  |
| 66 | negative | negative | 55 | other | yes | March 2020 |  |
| 66 | negative | negative | 48 | parent | no |  |  |
| 66 | negative | negative | 54 | parent | yes | March 2020 |  |
| 68 | positive | positive | 9 | screening positive | no |  | parent |
| 68 | negative | negative | 7 | sibling | no |  |  |
| 68 | positive | positive | 40 | parent | yes | March 2020 |  |
| 68 | positive | positive | 41 | parent | yes | March 2020 |  |
| 69 | positive | positive | 17 | screening positive | yes | March 2020 | NA |
| 69 | positive | positive | 54 | parent | yes | March 2020 |  |
| 70 | positive | positive | 4 | screening positive | yes | March 2020 | parent |
| 70 | negative | negative | 5 | sibling | no |  |  |
| 70 | positive | Positiv | 45 | parent | yes | March 2020 |  |
| 70 | positive | Positiv | 45 | parent | yes | March 2020 |  |
| 72 | positive | positive | 13 | screening positive | no |  | parent |
| 72 | negative | negative | 49 | parent | yes | March 2020 |  |
| 73 | positive | positive | 15 | screening positive | yes | March 2020 | screening positive |
| 73 | positive | positive | 41 | parent | yes | March 2020 |  |
| 73 |  |  | 48 | parent | yes | March 2020 |  |
| 76 | positive | positive | 13 | screening positive | yes | March 2020 | parent |
| 76 | positive | positive | 9 | screening positive | yes |  |  |
| 76 | negative | negative | 16 | sibling | yes |  |  |
| 76 | positive | positive | 18 | sibling | yes |  |  |
| 76 | positive | positive | 49 | parent | yes | March 2020 |  |
| 76 | negative | positive | 47 | parent | yes |  |  |
| 84 | positive | positive | 12 | screening positive | yes |  | parent |
| 84 | negative | positive | 9 | screening positive | no |  |  |
| 84 | positive | positive | 49 | parent | yes |  |  |
| 84 | positive | positive | 48 | parent | yes | March 2020 |  |
| 89 | positive | positive | 9 | screening positive | yes | March 2020 | screening positive |
| 89 | negative | negative | 6 | sibling | yes |  |  |
| 89 | negative | negative | 43 | parent | yes |  |  |
| 89 | negative | negative | 42 | parent | yes |  |  |
| 107 | positive | positive | 16 | screening positive | yes | March 2020 | screening positive |
| 107 | negative | negative | 6 | sibling | no |  |  |
| 107 | positive | negative | 8 | sibling | no |  |  |
| 107 | positive | positive | 46 | parent | yes | March 2020 |  |
| 107 | positive | positive | 46 | parent | yes | March 2020 |  |
| 108 | positive | positive | 10 | screening positive | yes | March 2020 | parent |
| 108 | negative | negative | 13 | sibling | no |  |  |
| 108 | negative | negative | 10 | sibling | no |  |  |
| 108 | positive | positive | 47 | parent | yes | March 2020 |  |
| 108 | negative | positive | 48 | parent | yes | March 2020 |  |
| 109 | positive | positive | 10 | screening positive | yes | March 2020 | parent |
| 109 | negative | negative | 50 | parent | NA |  |  |
| 109 | positive | positive | 46 | parent | yes | March 2020 |  |
| 110 | positive | positive | 14 | screening positive | yes | March 2020 | parent |
| 110 | positive | positive | 16 | sibling | yes |  |  |
| 110 | positive | positive | 4 | sibling | yes |  |  |
| 110 | positive | positive | 38 | parent | yes | March 2020 |  |
| 110 | positive | positive | 35 | parent | yes | March 2020 |  |
| 111 | positive | positive | 10 | screening positive | no |  | parent |
| 111 | negative | negative | 12 | sibling | no |  |  |
| 111 | negative | negative | 74 | grandparent | no |  |  |
| 111 | negative | positive | 51 | parent | yes |  |  |
| 111 | negative | negative | 78 | grandparent | no |  |  |
| 111 | negative | positive | 48 | parent | yes | March 2020 |  |
| 112 | positive | positive | 9 | screening positive | no |  | parent |
| 112* | positive | positive | 6 | screening positive | yes | March 2020 |  |
| 112 | negative | negative | 42 | parent | no |  |  |
| 112 | positive | positive | 43 | parent | yes | March 2020 |  |
| 113 | positive | positive | 4 | screening positive | yes |  | parent |
| 113 | positive | positive | 3 | sibling | no |  |  |
| 113 | positive | positive | 38 | parent | yes | March 2020 |  |
| 113 | negative | positive | 34 | parent | yes | March 2020 |  |
| 114 | positive | positive | 16 | screening positive | yes | March 2020 | parent |
| 114 | positive | positive | 13 | screening positive | yes | March 2020 |  |
| 114 | positive | positive | 11 | screening positive | no |  |  |
| 114 | negative | positive | 52 | parent | yes | March 2020 |  |
| 114 | positive | positive | 47 | parent | yes | March 2020 |  |

NA: not available

**Figure S1. Seroconversion rate distribution across age in children under 18 years of age.**

The black dots indicate the seroconversion rate per age. The calculated logistic regression model and corresponding 95% CI are shown (blue line and grey area). The probability of seroconversion increases with an odds ratio of 1·11 (95 % CI: 1·03-1·21, P=0·009) per year of age. For the 3 different age groups (0 - <6, 6 - <12 and 12 - <18 years), the mean seroconversion rates and respective 95% CI are depicted above the logistic regression model. Included are 4657 children that participated in the screening phase, including serologic testing. Seroconversion is defined based on positivity for both SARS-CoV-2 anti-spike IgG and anti-nucleocapsid IgA/IgG/IgM.

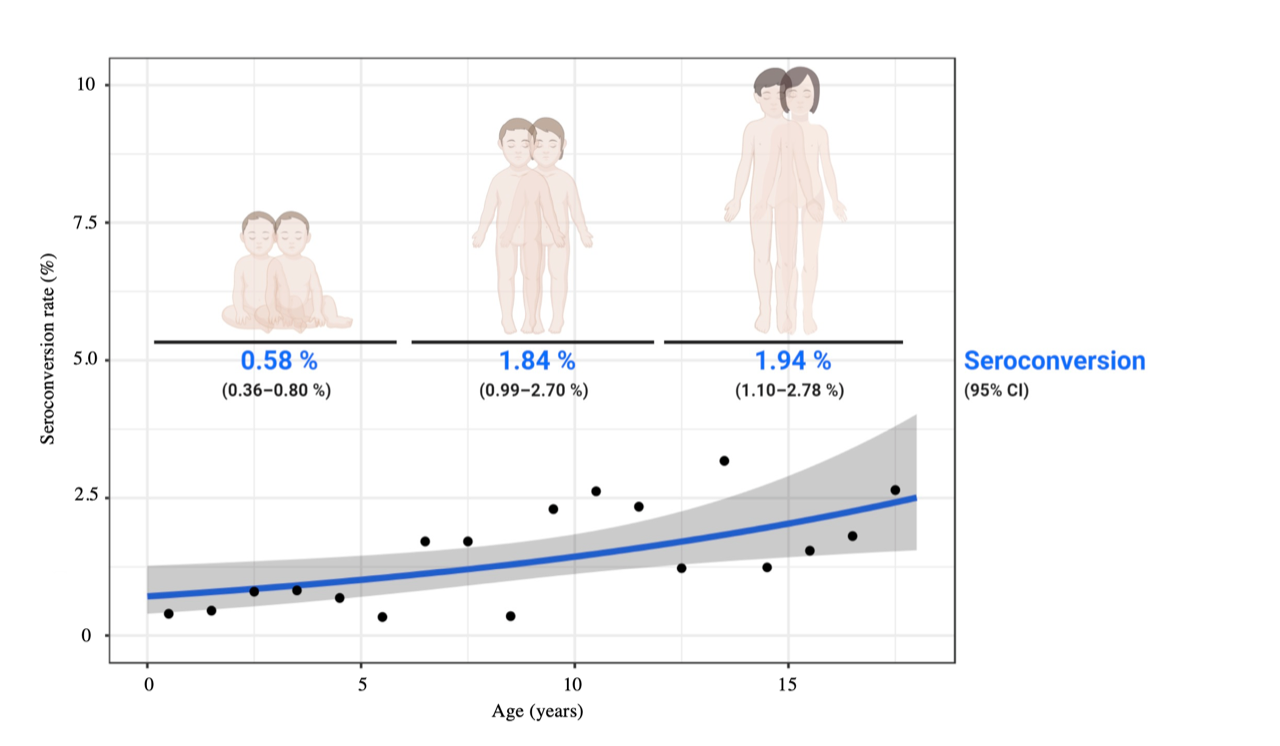

**Figure S2. Cumulative incidence per 100,000 in the general population of the city of Hamburg**

Data taken from daily situation reports of the Robert Koch Institute (https://www.rki.de/DE/Content/InfAZ/N/Neuartiges_Coronavirus/Situationsberichte/Gesamt.html), before, during, and after the screening phase of the C19.CHILD study. For raw data see also Supplementary Table S5.

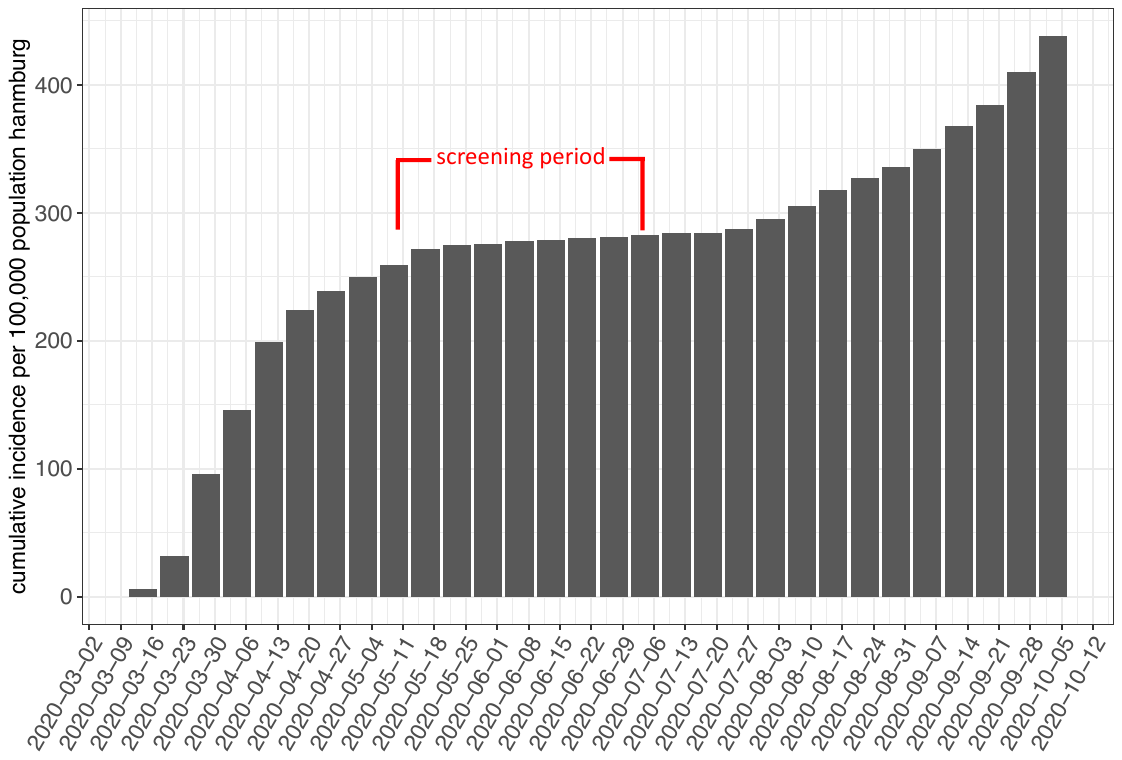
